# Reducing STI Burden in MSM with Doxy-PEP: Evidence from Individual-Based Modelling in Australia

**DOI:** 10.1101/2025.06.24.25330172

**Authors:** Ben Hui, Nicholas Medland, Nathan Ryder, Hayley Wareing, David Regan, Richard T. Gray

## Abstract

**Objectives:** Syphilis, gonorrhoea, and chlamydia are commonly diagnosed sexually transmitted infections (STIs) among men who have sex with men (MSM). Doxycycline post-exposure prophylaxis (Doxy-PEP) could be an effective public health intervention to reduce STI incidence. In this modelling study, we evaluate potential implementation strategies for Doxy-PEP roll-out to Australian MSM to inform clinical guidelines.

**Methods:** An individual-based mathematical model was developed to simulate the transmission of syphilis, gonorrhoea, and chlamydia within an urban MSM population in Australia. Individuals in the model form and dissolve regular and casual partnerships at rates based on publicly available sexual behaviour data, with infections transmitted through sexual contact within these partnerships. The impact on STI incidence over five- and ten-year periods was evaluated under different Doxy-PEP eligibility criteria, including HIV infection status and/or STI diagnosis history.

**Results:** Offering Doxy-PEP to individuals living with HIV and to current HIV pre-exposure prophylaxis (PrEP) users, or those with more than one positive STI diagnosis in the previous 12 months is estimated to reduce syphilis incidence by over 50% within 5 years. The incidence of gonorrhoea and chlamydia is predicted to decrease by over 40%, but the reduction in gonorrhoea incidence diminishes to less than 10% if Doxy-PEP efficacy against gonorrhoea declines over time due to increasing antimicrobial resistance.

**Conclusions:** Doxy-PEP could significantly reduce STI incidence, with the greatest impact observed for syphilis. However, the impact on gonorrhoea incidence may not be sustainable if Doxy-PEP efficacy wanes. Ongoing monitoring of Doxy-PEP efficacy and adherence is critical for reductions in STI incidence to be sustained.

**Key messages:** **What is already known on this topic**

Clinical trials have shown that doxycycline post-exposure prophylaxis (Doxy-PEP) can reduce the risk of acquiring bacterial STIs such as syphilis, gonorrhoea, and chlamydia. Given the potential resurgence of STIs among priority populations in Australia, the NSW Ministry of Health has considered including Doxy-PEP as part of its response to STIs. This study uses mathematical modelling to assess the potential epidemiological impact of prescribing Doxy-PEP to men who have sex with men (MSM) following a STI diagnosis.

**What this study adds**

The modelling suggests that introducing Doxy-PEP based on HIV status/HIV PrEP usage, or STI diagnosis could reduce syphilis and chlamydia incidence by over 40% within five years. A similar reduction in gonorrhoea incidence may also be possible; however, the impact could be limited to just 8% if antimicrobial resistance to doxycycline is already present or emerges because of the intervention.

**How this study might affect research, practice or policy**

Implementing Doxy-PEP alongside STI testing could reduce syphilis incidence among MSM in Australia. Similar reductions in other STIs may also be achievable; however, long-term effectiveness may be threatened by the emergence of doxycycline resistance associated with widespread Doxy-PEP use. To sustain these benefits, high uptake and adherence to Doxy-PEP, along with robust monitoring for antimicrobial resistance, will be essential.

## Background

Syphilis, caused by the bacterium Treponema pallidum, is a sexually transmissible infection (STI) that can lead to severe health complications, including cardiovascular and neurological disease, if left untreated (1). An estimated 8 million adults aged 15–49 acquired syphilis globally in 2022, with a resurgence in cases since 2013 among men who have sex with men (MSM) (2, 3, 4, 5). While syndromic management, regular screening, and antibiotic treatment remain the standard control measures for syphilis infection (1, 6, 7), the recent resurgence of syphilis has prompted the exploration of additional biomedical prevention strategies.

One potential strategy is the use of doxycycline as post-exposure antibiotic prophylaxis (Doxy-PEP) (8, 9). This involves taking a dose (usually 200 mg) of the antibiotic doxycycline up to 72 hours following condomless sexual exposure to reduce the risk of acquiring a bacterial STI. The efficacy of Doxy-PEP has been demonstrated in clinical trials, with the risk of first-episode syphilis infection reduced by up to 77% (10, 11, 12). These trials also reported varying efficacies in preventing gonorrhoea and chlamydia infections (10, 11, 12, 13, 14). Doxy-PEP is also highly acceptable to MSM (15, 16, 17). These findings suggest Doxy-PEP could have a large impact on syphilis epidemics among MSM, however, there are concerns about the potential for the development and spread of antimicrobial resistance, particularly in the case of gonorrhoea (9, 18). Evidence based clinical guidelines for the use of Doxy-PEP that consider its potential benefits and risks are required to inform implementation and maximise the public health benefits.

Several modelling studies have explored the potential impact of introducing Doxy-PEP on syphilis and/or STI incidence (19, 20, 21). An analysis using electronic health records in the United States has suggested that prescribing Doxy-PEP to gay and bisexual man, transgender women and non-binary people assigned male at birth would have averted 71% of STI diagnoses (19). Another study using United States data estimated that if 20% of MSM used Doxy-Pep with 80% adherence (defined as continuous use of Doxy-PEP throughout the entire simulation) syphilis incidence would decrease by 10% in ten years (21). A European based analysis estimated a risk reduction of 96% among MSM for doxycycline-based prevention strategies, as opposed to 59% for a condom-based approach (20). Additionally other studies have demonstrated real world effectiveness in both clinical cohorts (22) and at the population level (23) following Doxy-PEP implementation.

Like other high-income countries, Australia has seen increasing trends in STIs and attributed harms, including congenital syphilis (4). In the most populous state of New South Wales (NSW), annual notifications of syphilis have tripled over the last decade, reaching a new maximum of 2,034 infectious syphilis notifications in 2023, with more than half of these cases (1,107) among MSM (24). The NSW STI Strategy 2022-2026 focuses primarily on syphilis, gonorrhoea, and chlamydia and aims to reduce the prevalence and impacts of these STIs in NSW (25). As part of the implementation of the strategy, the NSW Ministry of Health is considering the evidence-based use of Doxy-PEP, as well as the likely impacts on clinical services and surveillance.

In this study we used mathematical modelling to evaluate the potential impact of Doxy-PEP on STIs among MSM in NSW. Our primary focus was on syphilis, but we also considered the impact on chlamydia and gonorrhoea, and the potential effects of widespread Doxy-PEP implementation on antibiotic resistance levels within the population. Resistance is a particular concern for gonorrhoea, as Neisseria gonorrhoeae strains with high-level tetracycline resistance are already widespread in Australia (26). This issue could worsen if, as a recently published study suggests, the rollout of Doxy-PEP becomes associated with an increase in tetracycline resistance (18).

Through engagement with the NSW Ministry of Health we simulated feasible Doxy-PEP implementation scenarios based on realistic eligibility criteria over a ten-year period to estimate the potential reduction in incidence of syphilis, gonorrhoea, and chlamydia. To achieve this, we developed an individual-based model (IBM) to simulate the transmission of syphilis, gonorrhoea, and chlamydia (referred to collectively here as STIs) among MSM in NSW, Australia. The model builds on previously developed and implemented models of STI transmission (27) and tracks the formation and dissolution of sexual partnerships within the model population and the transmission of STIs within them.

## Methods

We developed an individual based model (IBM) of 104 000 gay and bisexual men in NSW. The model based on a previously developed model of gonorrhoea transmission, extended to track the transmission of syphilis, chlamydia, and gonorrhoea simultaneously (28). The primary reason for adopting an IBM for this analysis is to have the ability to track STI incidence and diagnosis history at the individual level. This capability allows for a detailed examination of practical scenarios, which can be directly tied to real-world implementation. Specifically, our model design facilitates the investigation of strategies aimed at individuals— such as offering Doxy-PEP at the time of an STI test and establishing Doxy-PEP eligibility based on STI diagnosis history. These approaches align with the selective use of Doxy-PEP as recommended by current consensus (9).

The technical details of the model are provided in the supplementary material. The model is implemented in Java (Standard Edition 11) and is openly accessible on GitHub under an open-access license: https://github.com/The-Kirby-Institute/Launcher_Doxy_PEP.

### Summary of model

Before introduction of STIs into the model, a partnership network was generated based on publicly available sexual behaviour data (29, 30). Subsequently, STIs were incorporated into the model, and simulations were conducted over a 10-year period, starting with the hypothetical introduction of Doxy-PEP in 2025. Note that while syphilis notifications in NSW and nationally reached their highest levels in 2022, the rate of increase has noticeably slowed during 2019–2022 compared to the rapid rise observed between 2017 and 2019 (5).

### Calibration

The model is calibrated using national incidence data for syphilis, gonorrhoea, and chlamydia from the 2023 Annual Surveillance Report (5). The calibration involved adjusting model parameters that describe anatomical site-specific transmission rates, durations of infection, and the probability of symptomatic infection. The process followed a multi-stage approach. First, prior distributions for each parameter were defined based on published sources and previous models. A simplex algorithm (31) was then employed to adjust the mean values of these distributions, ensuring that the STI incidence aligns with the range observed in the national data. Additional parameter sets were then resampled from the priors for the simulation run. The top 1,000 simulations that most closely aligned with the upward syphilis incidence trends from 2018 to 2021 were selected as the baseline model. Finally, various scenarios outlining different Doxy-PEP implementation strategies were applied to the selected simulations, and the resulting changes in STI incidence are reported.

Our model projects that syphilis incidence will plateau by 2025 as recent data suggest that syphilis incidence is likely to rise only slowly or stabilise after 2022 (32). This projection is further supported by the latest national surveillance data from 2024, which show a continued decline in syphilis incidence among both HIV-negative and HIV-positive gay and bisexual men since 2021(5).

### Scenarios simulated

Scenarios were developed based on four Doxy-PEP eligibility criteria (summarised in **Error! Reference source not found.**) considered in Traeger et al (33). Eligibility for Doxy-PEP is based on STI diagnosis (syphilis diagnosis only for Scenario 2, and syphilis, gonorrhoea or chlamydia diagnosis for Scenario 3 and 4), or if the individual is living with HIV or using HIV pre-exposure prophylaxis (HIV PrEP) (Scenario 1 and 4).

**Table 1:** Doxy-PEP eligibility scenarios investigated in this study. These specify the eligibility criteria for individuals to be offered Doxy-PEP at the time of testing for STIs.

| # | Doxy-PEP eligibility scenarios |
| --- | --- |
| 1 | Living with HIV or currently an HIV PrEP user |
| 2 | Have a positive syphilis diagnosis at the time of testing |
| 3 | Have a positive STI (syphilis, gonorrhoea or chlamydia) diagnosis at the time of testing, and have had at least 1 more positive STI diagnosis in the last 12 months |
| 4 | Any of Scenarios 1, 2, or 3 |

For each scenario, we assumed that on presentation for STI testing, eligible individuals will be offered a prescription that provides access to Doxy-PEP for 180 days. After 180 days, they can renew the prescription as long as they remain eligible. In our baseline analysis, we consider an ideal situation where the preventative efficacy of Doxy-PEP remains constant (77%, 22%, and 78% for syphilis, gonorrhoea, and chlamydia infections, respectively) throughout the entire simulation. We also assumed that Doxy-PEP is used by all eligible individuals (i.e., uptake of 100%, defined as the percentage of eligible individuals who commence Doxy-PEP upon STI diagnosis) following all sexual encounters (i.e., adherence of 100%, defined as the probability that an individual will use Doxy-PEP following a sexual encounter, as a percentage of sexual encounters) for the entirety of the prescription (i.e., duration of use of 180 days, defined as the duration of time an individual continues to use Doxy-PEP following receipt of a prescription).

Our ideal implementation assumes that all individuals eligible for Doxy-PEP will initiate and fully adhere to the recommended regimen. To assess the implications of non-ideal implementation, we simulated a scenario under non-ideal implementation based on Scenario 3 (i.e., eligibility based on STI diagnosis), where uptake and adherence are less than 100% or the duration of use is reduced.

### Impact on gonorrhoea drug resistance

To estimate the potential impact of selective pressure on doxycycline-sensitive gonorrhoea infections following the rollout of Doxy-PEP, we explored scenarios where Doxy-PEP efficacy is infection-specific. In this analysis, gonorrhoea infections are classified as either Doxy-PEP-sensitive or Doxy-PEP-resistant. For simplicity, we assumed a conservative scenario where both categories of infections have identical characteristics (e.g., transmission probabilities and durations), except for Doxy-PEP’s capacity to prevent infection. Doxy-PEP efficacy is assumed to be 1 for Doxy-PEP-sensitive infections and 0 for Doxy-PEP-resistant infections. These categories are considered stable through transmission (i.e., an infection caused by a Doxy-PEP-resistant strain will also be Doxy-PEP-resistant until treatment or recovery). Furthermore, we assumed the baseline Doxy-PEP efficacy for gonorrhoea corresponds to the proportion of gonorrhoea infections that are Doxy-PEP-sensitive at the time of Doxy-PEP rollout. Based on this assumption, 22% of gonorrhoea cases were considered Doxy-PEP-sensitive prior to the introduction of Doxy-PEP.

For each scenario we ran 1,000 simulations. Gonorrhoea incidence at 5 and 10 years are compared with the corresponding simulation in the absence of Doxy-PEP, with the median and interquartile range (IQR) of the difference across the simulations reported.

## Results

### Infectious syphilis incidence in the absence of Doxy-PEP

Syphilis transmission in our model between 2017-2030 under the status quo scenario is shown in Figure S2-1 in the supplementary material. In the 2017-2021 period, the infectious syphilis incidence rate was estimated to rise from 3.4 per 100 person-years (PY) (IQR: 3.0 – 3.7) to 5.5 per 100 PY (IQR: 5.0 – 6.0). The syphilis incidence rate is then projected to stabilise, peaking at 6.2 per 100 PY (IQR: 5.6 – 6.6) in 2025 before dropping slightly to 5.9 per 100 PY (IQR: 5.4 – 6.3) by 2030.

### Infectious syphilis incidence under Doxy-PEP

For all scenarios involving Doxy-PEP, we assume it is introduced in 2025, when the modelled syphilis incidence rate is 6.2 per 100 PY (IQR: 5.6-6.6). Without Doxy-PEP, the predicted incidence rates in the 5^th^ year following its hypothetical introduction in 2025, are 5.9 (5.4-6.3) per 100 PY for syphilis, 28.8 (23.7-33.7) per 100 PY for gonorrhoea, and 31.0 (30.5-31.5) per 100 PY for chlamydia. By the 10^th^ year, the incidence rates are 5.4 (5.0-5.8) per 100 PY for syphilis, 28.5 (23.4-33.6) per 100 PY for gonorrhoea, and 31.0 (30.5-31.5) per 100 PY for chlamydia.

### Fixed Doxy-PEP efficacy

The results in Figure 1 show STI incidence for the modelled population over 10 years under the ideal implementation scenario, assuming constant Doxy-PEP efficacy of 77% for syphilis (top left), 22% for gonorrhoea (bottom left), and 78% for chlamydia (top right). Our model predicts a substantial reduction in infectious syphilis incidence across all Doxy-PEP eligibility criteria considered. Gonorrhoea and chlamydia incidence is also predicted to decrease significantly under all scenarios except Scenario 2, where Doxy-PEP is offered only to those who test positive for syphilis. In this scenario, only a small reduction in incidence is expected for gonorrhoea and chlamydia, with a more substantial reduction for syphilis.

**Figure 1:**
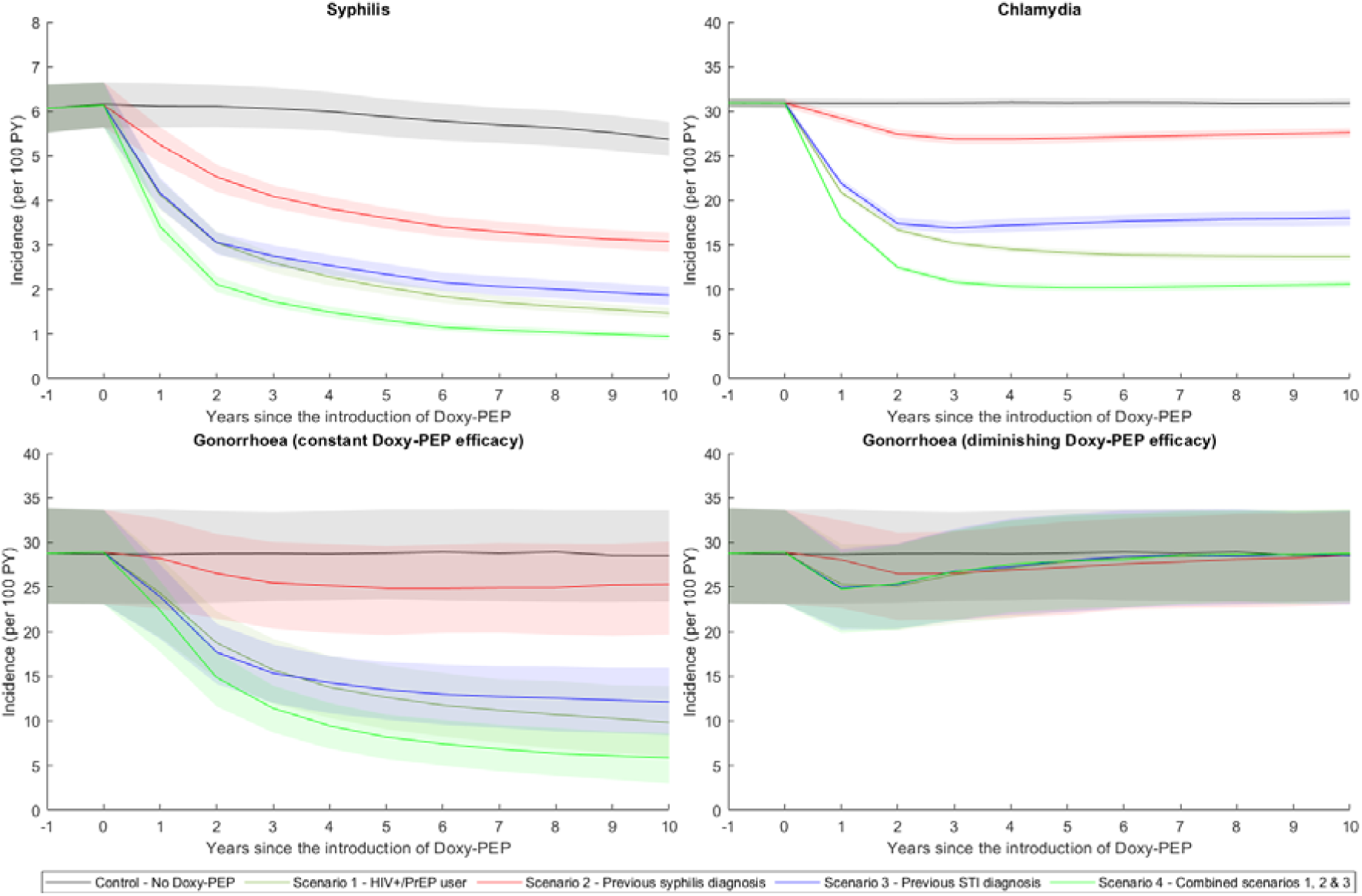
Projected Impact of Doxy-PEP on STI Incidence. This figure presents the projected impact of Doxy-PEP on the incidence of four sexually transmitted infections over time, across various eligibility scenarios. Panels show syphilis (top left), chlamydia (top right), gonorrhoea assuming constant population-wide Doxy-PEP efficacy (bottom left), and gonorrhoea assuming diminishing efficacy due to selective pressure (bottom right). All scenarios are compared against a status quo without Doxy-PEP (black line). Solid lines represent the median incidence across 1,000 simulations, while shaded areas indicate the interquartile range for each scenario.

**Error! Reference source not found.** shows the relative reduction in the cumulative number of STI infections due to the implementation of Doxy-PEP strategies, compared to the cumulative number of infections over the same period without Doxy-PEP. For example, consider the scenario where Doxy-PEP is available to individuals with more than one STI diagnosis in the last 12 months (Scenario 3 in **Error! Reference source not found.**). Under ideal implementation, this strategy could reduce syphilis infections by 50.6% by the fifth year (with the mean incidence rate dropping from 5.9 to 2.3 per 100 PY) relative to the scenario without Doxy-PEP. By the tenth year, this reduction deepens to 57.1% (with the mean incidence rate falling from 5.4 to 1.9 per 100 PY).

**Table 2:**
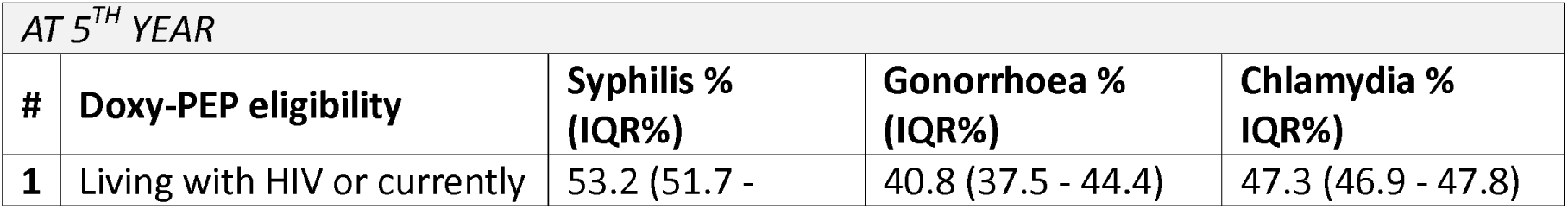
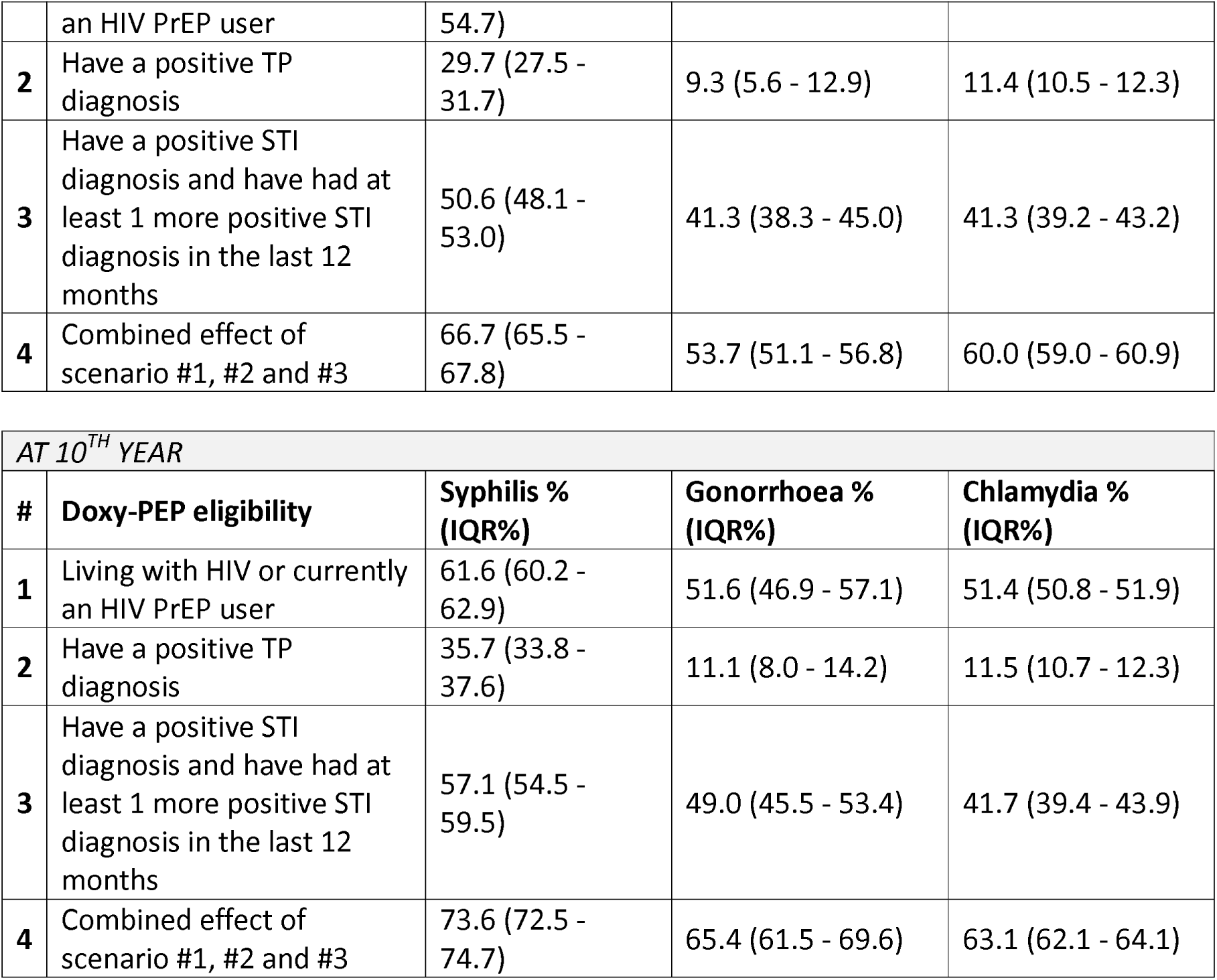
Estimated percentage reduction in cumulative STI infections at 5 and 10 years (2030 and 2035) after Doxy-PEP introduction, relative to a scenario without Doxy-PEP. The estimates are based on the scenarios outlined in ***Error! Reference source not found.***, assuming ideal uptake, duration of use, and adherence, with Doxy-PEP efficacy remains constant throughout simulation. The values represent the median and the interquartile range (in parentheses) of the percentage reduction for each STI compared to the without Doxy-PEP scenario.

### Population-wide efficacy of Doxy-PEP diminish over time due to antibiotic selective pressure

The results shown in the bottom right of Figure 1 and **Error! Reference source not found.** show NG incidence for the modelled population over 10 years under ideal implementation, assuming population-wide Doxy-PEP efficacy decreases over time due to selective pressure (in contrast to the fixed population-wide efficacy modelled previously). The result show reductions in gonorrhoea incidence cannot be maintained. Specifically, gonorrhoea incidence experiences a rebound well within two years or less.

This rebound in STI incidence is linked to the declining proportion of infections that remain sensitive to Doxy-PEP (see **Error! Reference source not found.**). Our results suggest that, except in Scenario 2, nearly all gonorrhoea infections are resistant to Doxy-PEP within 5 years of its introduction.

**Table 3:** Estimated percentage reduction in cumulative gonorrhoea infections and the percentage of infections remaining susceptible to Doxy-PEP at 5 and 10 years (2030 and 2035) post-Doxy-PEP introduction, relative to a scenario without Doxy-PEP. The estimates are based on the scenarios outlined in ***Error! Reference source not found.***, assuming ideal uptake, duration of use, and adherence. This model assumes population-wide Doxy-PEP efficacy varying over time due to selective pressure. The values represent the median and the interquartile range (in parentheses) of the percentage reduction for gonorrhoea compared to the status quo (i.e., without Doxy-PEP).

| AT 5<sup>TH</sup> YEAR |  |  |  |
| --- | --- | --- | --- |
| # | Doxy-PEP eligibility | Percentage of gonorrhoea | Percentage of gonorrhoea |

|  |  | **incidence averted**<br>**compared to no Doxy-PEP**<br>**(IQR%)** | **infections remaining**<br>**susceptible to Doxy-PEP**<br>**(IQR%)** |
| --- | --- | --- | --- |
| **1** | Living with HIV or currently an HIV PrEP user | 8.1 (4.1 - 12.1) | 0.0 (0.0 - 0.0) |
| **2** | Have a positive TP diagnosis | 5.9 (1.5 - 10.3) | 2.3 (1.3 - 3.5) |
| **3** | Have a positive STI diagnosis and have had at least 1 more positive STI diagnosis in the last 12 months | 7.7 (3.8 - 11.8) | 0.0 (0.0 - 0.0) |
| **4** | Combined effect of scenario #1, #2 and #3 | 8.1 (4.1 - 12.3) | 0.0 (0.0 - 0.0) |

| *AT 10<sup>TH</sup> YEAR* |  |  |  |
| --- | --- | --- | --- |
| **#** | **Doxy-PEP eligibility** | **Percentage of gonorrhoea**<br>**incidence averted**<br>**compared to no Doxy-PEP**<br>**(IQR%)** | **Percentage of gonorrhoea**<br>**infections remaining**<br>**susceptible to Doxy-PEP**<br>**(IQR%)** |
| **1** | Living with HIV or currently an HIV PrEP user | 4.6 (1.2 - 7.8) | 0.0 (0.0 - 0.0) |
| **2** | Have a positive TP diagnosis | 3.9 (0.6 - 7.8) | 0.0 (0.0 - 0.4) |
| **3** | Have a positive STI diagnosis and have had at least 1 more positive STI diagnosis in the last 12 months | 4.2 (1.2 - 7.8) | 0.0 (0.0 - 0.0) |
| **4** | Combined effect of scenario #1, #2 and #3 | 4.3 (1.2 - 8.0) | 0.0 (0.0 - 0.0) |

### Effects of reduced duration, adherence, and uptake

We explored the potential impact of reduced duration of use (the time an individual continues to use Doxy-PEP after receiving a prescription) from 180 days to 90 days, and reduced adherence (the probability an individual uses Doxy-PEP after a sexual encounter) from 100% to 75%. Under these conditions, the relative reduction in cumulative syphilis incidence by the tenth year is expected to drop from 57.1% to 40.7% (see Table S2-1 in the supplementary material). A lower uptake rate results in the relative reduction in cumulative syphilis incidence at 10th year declining to 46.1% for a 50% uptake rate, and 33.5% for a 25% uptake rate (see Table S2-2 in the supplementary material).

## Discussion

Our modelling study estimated the potential impact of Doxy-PEP on STI incidence among MSM in NSW, Australia. While the proposed intervention primarily focused on syphilis, we also assessed the effects of Doxy-PEP on gonorrhoea, chlamydia, and the prevalence of gonococcal resistance in the population. By designing simulation scenarios based on feasible eligibility criteria, our findings provide guidance for the development of Doxy-PEP implementation strategies that balance reducing STI incidence with responsible antibiotic prescribing and use. These results highlight Doxy-PEP’s potential to contribute significantly to achieving syphilis control.

The model predicts that the provision of Doxy-PEP for eligible MSM, under ideal conditions and assuming stable levels of doxycycline resistance in the population, could result in substantial reductions in the incidence of syphilis, gonorrhoea, and chlamydia. Specifically, if all eligible individuals with a positive syphilis diagnosis initiate Doxy-PEP and adhere fully to the recommended regimen, syphilis incidence could be reduced by 30% within five years.

This reduction exceeds 50% if eligibility is expanded to include people living with HIV, people who are using HIV PrEP, or individuals with a positive STI diagnosis. Our modelling approach and results are similar to and consistent with that of Tran et al. (21). Their model predicted a 55% reduction in syphilis incidence over 10 years with 100% uptake and adherence consistent with our findings, despite differences in the interpretation of uptake (where Tran et al. defined uptake at the population level, compared to uptake among individuals participating in STI testing).

If Doxy-PEP uptake among eligible individuals declines over time, achieving substantial reductions in STI incidence becomes increasingly challenging. For instance, a 44% decrease in incidence over five years necessitates at least 75% of Doxy-PEP users to continue their prescriptions consistently. This outcome emphasises that while offering Doxy-PEP through STI screening can significantly reduce syphilis incidence in the short-term, maintaining this reduction requires continued Doxy-PEP use through regular prescription renewals, or alternatively, broader rollout strategies not solely based on STI diagnoses, such as awareness campaigns that promote ongoing Doxy-PEP use. Further research is needed to establish realistic targets for Doxy-PEP uptake, especially given the potential challenges, such as maintaining adherence as syphilis incidence declines.

The impact of antibiotic resistance in STIs also needs to be considered. Our modelling indicates that, due to selective pressure from the Doxy-PEP rollout, almost all gonorrhoea infections in the population could become Doxy-PEP-resistant within five years. Since doxycycline is not the recommended treatment for gonorrhoea in Australia the direct significance of this may be low. However emerging data suggest co-selection of dual doxycycline and ceftriaxone resistant strain may occur with far greater clinical and public health significance (34). While these projections may reflect overly cautious assumptions (such as both Doxy-PEP-sensitive and resistant infections sharing similar transmission characteristics), the rapid emergence of resistance is not unrealistic. A study based on surveillance data from MSM in King County in United States (a similar setting to NSW) reported a rise in tetracycline resistance Neisseria gonorrhoeae strain from 27% to 70% within just one year of Doxy-PEP rollout (18). If this rise in resistance corresponds to a reduction in Doxy-PEP efficacy, it suggests that the impact of Doxy-PEP on gonorrhoea may not be sustainable.

While previous modelling studies have examined the impact of Doxy-PEP on STI incidence under various scenarios (19, 20, 21), our model offers a novel approach by incorporating the transmission dynamics and the impact of incidence across multiple STIs, while evaluating feasible implementation strategies based on current STI control measures. Specifically, we explored the introduction of Doxy-PEP through STI diagnosis, HIV status, and HIV PrEP usage— approaches considered feasible by NSW Health. This framework allows for a more thorough evaluation of implementation strategies, including the concurrent protection Doxy-PEP offers against multiple STIs, the diminishing impact on incidence as the number of STI diagnoses decreases, and the potential risk of antibiotic resistance from selective pressure.

Our modelling study has several limitations. Regarding STI transmission, the model relies on natural history parameters that are often uncertain and based on historical studies. Data concerning anatomical site-specific transmission, in particular, remain limited. Consequently, we cannot exclude the possibility that different parameter values might produce similar prevalence and/or incidence figures within the model (35). The model also did not include HIV transmission dynamics. It instead assumes a static proportion of the population are living with HIV or using HIV PrEP as determined by partner acquisition and testing behaviour. However, including HIV transmission in our study would have reduced the model’s tractability while having minimal impact on the study findings, as HIV transmission rates in Australia and NSW are low relative to other STIs and continue to decline (5, 36). Additionally, due to a lack of suitable data, the model assumed that STI transmissions occurred independently. It did not account for potential biological interactions (e.g., altered transmission probabilities) or behavioural interactions (e.g., how symptoms of multiple STIs might influence diagnosis and testing behaviours) arising from co-infections. Beyond STI transmission, we assumed that most model parameters remain constant throughout the simulations. Specifically, we assumed sexual behaviour characteristics, such as the number of partnerships, frequency of unprotected sex acts, and the STI testing rate, remain unchanged by Doxy-PEP availability. The probability of using Doxy-PEP is also modelled as being the same at the individual level, whereas in practice, usage may depend on the risk associated with specific events or partners. These simplifying assumptions are necessary due to the limited data on how current Doxy-PEP usage might influence sexual behaviour changes. Finally, while our results suggest that the rollout of Doxy-PEP could lead to the emergence of antimicrobial resistance, it is unclear whether this is directly applicable in our context, given that high-level tetracycline resistance is already widespread among MSM prior to the Doxy-PEP rollout. Specifically, our assumptions—that Doxy-PEP-resistant strains share the same natural history as Doxy-PEP-sensitive strains, and that the efficacy of Doxy-PEP at the start of rollout corresponds to the prevalence of resistant strains in the population—may be overly optimistic and could lead to an overestimation of the emergence rate. Nevertheless, since other research indicates a rapid increase in gonorrhoea resistance following Doxy-PEP rollout (18), the potential risk of resistance emergence should still be carefully considered.

In summary, our modelling indicates that introducing Doxy-PEP through STI testing could lead to notable reduction in syphilis incidence among MSM in Australia, with sustained long-term reductions achievable if population-level Doxy-PEP usage is maintained. While Doxy-PEP may also reduce the incidence of other STIs including gonorrhoea and chlamydia, these benefits are likely to be unsustainable should its population-wide effectiveness wanes over time due to factors like non-adherence and, specifically for gonorrhoea, the emergence of antibiotic resistance. Continuous monitoring of Doxy-PEP adherence and population-level efficacy will be crucial to ensuring the long-term success of syphilis reduction efforts.

## Data Availability

The modelling code is available online thru GitHub
All other data produced in the present study are available upon reasonable request to the authors

https://github.com/The-Kirby-Institute/Launcher_Doxy_PEP

## Acknowledgments

This project was supported by the BBV & STI Research, Intervention and Strategic Evaluation (BRISE) program, funded by the NSW Ministry of Health. The Kirby Institute is funded by the Australian Government Department of Health and Aged Care. The views expressed in this publication do not necessarily represent the position of the Australian Government

This research was produced in whole or part by UNSW Sydney researchers and is subject to the UNSW intellectual property policy. For the purposes of Open Access, the author has applied a Creative Commons Attribution CC-BY public copyright licence to any Author Accepted Manuscript (AAM) version arising from this submission.

## References

1. National Health Service (NHS). Syphilis 2022 [Available from: https://www.nhs.uk/conditions/syphilis/.

2. World Health Organization. Syphilis 2024 [Available from: https://www.who.int/news-room/fact-sheets/detail/syphilis.

3. Fountain H, Charles H, Simms I, Migchelsen SJ, Mohammed H, Sinka K. Tracking the syphilis epidemic in England: 2013 to 2023. London: UK Health Security Agency; 2024.

4. Phua G, White C. The resurgence of syphilis in Australia. Aust J Gen Pract. 2024;53(3):133–7.

5. King J, Kwon JA, McManus H, Gray R, McGregor S. HIV, viral hepatitis and sexually transmissible infections in Australia: Annual surveillance report 2024. Sydney: Kirby Insitute, UNSW Sydney; 2024.

6. Centers for Disease Control and Prevention. Syphilis 2024 [Available from: https://www.cdc.gov/std/treatment-guidelines/syphilis.htm.

7. ASHM. Syphilis 2024 [Available from: https://sti.guidelines.org.au/sexually-transmissible-infections/syphilis/.

8. Meyerowitz EA, Liang R, Bishop D, Mullis CE. Put a little doxy-PEP in your step: Using doxycycline to prevent chlamydia, syphilis, and gonorrhea infections. PLoS Pathog. 2024;20(9):e1012575.

9. Cornelisse VJ, Riley B, Medland NA. Australian consensus statement on doxycycline post-exposure prophylaxis (doxy-PEP) for the prevention of syphilis, chlamydia and gonorrhoea among gay, bisexual and other men who have sex with men. Med J Aust. 2024;220(7):381–6.

10. Sokoll PR, Migliavaca CB, Doring S, Traub U, Stark K, Sardeli AV. Efficacy of postexposure prophylaxis with doxycycline (Doxy-PEP) in reducing sexually transmitted infections: a systematic review and meta-analysis. Sex Transm Infect. 2024.

11. Luetkemeyer AF, Donnell D, Dombrowski JC, Cohen S, Grabow C, Brown CE, et al. Postexposure Doxycycline to Prevent Bacterial Sexually Transmitted Infections. N Engl J Med. 2023;388(14):1296–306.

12. Molina JM, Charreau I, Chidiac C, Pialoux G, Cua E, Delaugerre C, et al. Post-exposure prophylaxis with doxycycline to prevent sexually transmitted infections in men who have sex with men: an open-label randomised substudy of the ANRS IPERGAY trial. Lancet Infect Dis. 2018;18(3):308–17.

13. Cannon CA, Celum CL. Doxycycline postexposure prophylaxis for prevention of sexually transmitted infections. Topics in antiviral medicine. 2023;31(5):566–75.

14. Cornelisse VJ, Ong JJ, Ryder N, Ooi C, Wong A, Kenchington P, et al. Interim position statement on doxycycline post-exposure prophylaxis (Doxy-PEP) for the prevention of bacterial sexually transmissible infections in Australia and Aotearoa New Zealand - the Australasian Society for HIV, Viral Hepatitis and Sexual Health Medicine (ASHM). Sex Health. 2023.

15. Spinelli MA, Scott HM, Vittinghoff E, Liu AY, Coleman K, Buchbinder SP. High Interest in Doxycycline for Sexually Transmitted Infection Postexposure Prophylaxis in a Multicity Survey of Men Who Have Sex With Men Using a Social Networking Application. Sex Transm Dis. 2019;46(4):e32–e4.

16. Holt M, Bavinton BR, Calabrese SK, Broady TR, Clackett S, Cornelisse VJ, et al. Acceptability of doxycycline prophylaxis, prior antibiotic use, and knowledge of antimicrobial resistance among Australian gay and bisexual men and non-binary people. Sex Transm Dis. 2024.

17. Chow EPF, Fairley CK. Use of doxycycline prophylaxis among gay and bisexual men in Melbourne. Lancet HIV. 2019;6(9):e568–e9.

18. Soge OO, Thibault CS, Cannon CA, McLaughlin SE, Menza TW, Dombrowski JC, et al. Potential Impact of Doxycycline Post-Exposure Prophylaxis on Tetracycline Resistance in Neisseria gonorrhoeae and Colonization with Tetracycline-Resistant Staphylococcus aureus and Group A Streptococcus. Clin Infect Dis. 2025.

19. Traeger MW, Mayer KH, Krakower DS, Gitin S, Jenness SM, Marcus JL. Potential impact of doxycycline post-exposure prophylaxis prescribing strategies on incidence of bacterial sexually transmitted infections. Clin Infect Dis. 2023.

20. Hahn A, Frickmann H, Loderstadt U. Modelling of doxycycline-based prevention of bacterial sexually transmitted infections in comparison to condom-based and test-based prevention. Eur J Microbiol Immunol (Bp). 2024;14(1):50–8.

21. Tran NK, Goldstein ND, Welles SL. Countering the rise of syphilis: A role for doxycycline post-exposure prophylaxis? Int J STD AIDS. 2022;33(1):18–30.

22. Traeger MW, Leyden WA, Volk JE, Silverberg MJ, Horberg MA, Davis TL, et al. Doxycycline Postexposure Prophylaxis and Bacterial Sexually Transmitted Infections Among Individuals Using HIV Preexposure Prophylaxis. Jama Intern Med. 2025;185(3):273–81.

23. Vanbaelen T, Manoharan-Basil SS, Kenyon C. Doxy-PEP could select for ceftriaxone resistance in Neisseria gonorrhoeae. Lancet Infect Dis. 2025.

24. NSW Health. NSW Sexually Transmissible Infections Data Report - January to December 2023. 2024.

25. New South Wales Health Centre for Population Health. NSW Sexually Transmissible Infections Strategy 2022-2026. 2022 [Available from: https://www.health.nsw.gov.au/sexualhealth/Pages/nsw-sti-strategy.aspx.

26. Lahra MM, Van Hal S, Hogan TR. Australian Gonococcal Surveillance Programme Annual Report, 2022. Commun Dis Intell (2018). 2023;47.

27. Hui BB, Ward JS, Guy R, Law MG, Gray RT, Regan DG. Impact of Testing Strategies to Combat a Major Syphilis Outbreak Among Australian Aboriginal and Torres Strait Islander Peoples: A Mathematical Modeling Study. Open Forum Infect Dis. 2022;9(5):ofac119.

28. Hui BB, Padeniya TN, Rebuli N, Gray RT, Wood JG, Donovan B, et al. A Gonococcal Vaccine Has the Potential to Rapidly Reduce the Incidence of Neisseria gonorrhoeae Infection Among Urban Men Who Have Sex With Men. J Infect Dis. 2022;225(6):983–93.

29. Second Australian Study of Health and Relationships (ASHR2). Sex Health. 2014;11(5):381–509.

30. GBQ+ Community Periodic Surveys. [Available from: https://www.unsw.edu.au/research/csrh/our-projects/gay-community-periodic-surveys.

31. Lagarias JC, Reeds JA, Wright MH, Wright PE. Convergence properties of the Nelder-Mead simplex method in low dimensions. Siam J Optimiz. 1998;9(1):112–47.

32. Traeger MW, Guy R, Taunton C, Chow EPF, Asselin J, Carter A, et al. Syphilis testing, incidence, and reinfection among gay and bisexual men in Australia over a decade spanning HIV PrEP implementation: an analysis of surveillance data from 2012 to 2022. The Lancet Regional Health – Western Pacific. 2024;51.

33. Traeger MW, Mayer KH, Krakower DS, Gitin S, Jenness SM, Marcus JL. Potential impact of doxycycline post-exposure prophylaxis prescribing strategies on incidence of bacterial sexually transmitted infections. Clin Infect Dis. 2023.

34. Kenyon C. Doxycycline post-exposure prophylaxis could theoretically select for resistance to various antimicrobials in 19 pathobionts: an in silico analysis. Int J Infect Dis. 2024;142:106974.

35. Spicknall IH, Mayer KH, Aral SO, Romero-Severson EO. Assessing uncertainty in an anatomical site-specific gonococcal infection transmission model of men who have sex with men. Sex Transm Dis. 2018.

36. NSW Health. NSW HIV Strategy 2021–2025 : 2023 Annual Data Report. 2023.

