## Supplementary material for "Reducing STI Burden in MSM with Doxy-PEP: Evidence from Individual-Based Modelling in Australia"

### Supplementary Material: Modelling the Impact of Doxycycline Post-Exposure Prophylaxis (Doxy-PEP) on STI Burden Among MSM in Australia

#### A1. Technical appendix

##### Overview

A stochastic individual-based mathematical model was developed to estimate the potential impact of Doxy-PEP on the incidence of STIs (syphilis, gonorrhoea and chlamydia) in men who have sex with men (MSM) in NSW. In this model, Doxy-PEP is made available to MSM at the time of presentation for STI testing, based on a range of eligibility criteria. The primary focus of the study is syphilis, but we also consider gonorrhoea and chlamydia because Doxy-PEP may also impact these.

This model simulates STI transmission among sexually active MSM, represented as individuals connected through a dynamic partnership network that evolves over time. The model's main output is the incidence of each STI, defined as the rate of new infections per person-year. A new infection for a specific STI occurs when a previously uninfected individual (at any anatomical site) becomes infected at least one anatomical site. The course of infection with an STI is independent of the infection status for other STIs. The median incidence and associated interquartile range, calculated over 1,000 simulations for each scenario evaluated, are reported in the Results section of the main text.

The modelled population consists of 104,000 MSM. Each individual can become infected with Treponema pallidum (TP, the causative bacterium of syphilis) during sexual contact with another individual who has infectious syphilis. Each individual also has three anatomical sites (penis, rectum, and pharynx) that can be infected, independently, with Neisseria gonorrhoeae (NG, the causative bacterium of gonorrhoea) and/or Chlamydia trachomatis (CT, the causative bacterium of chlamydia) upon sexual contact with another infectious individual. The infection status for each individual is updated daily throughout each simulation run.

In this model, sexual contact occurs within partnerships. The duration of these partnerships can range from a single day in casual partnerships to an average of four years in regular partnerships. Before STIs are introduced to the model, dynamic partnership networks are generated, and the same network is utilised across different scenarios to enable comparison of Doxy-PEP implementation strategies. See section 0 below for a further explanation of the network generation process and the parameters involved.

##### Partnership network

A separate process, independent of STI transmission, is used to generate the dynamic network that describes partner formation and dissolution within each simulation run. Conceptually, this network is represented by a graph, with individuals as vertices and partnerships as edges. Specific parameters describe the time duration for a partnership (edge) and when it starts and stops. Two individuals in the network can have more than one partner, as long as the partnership periods do not overlap. All parameters used in network generation are listed and defined in Table 1-1. Ten networks were generated for this study and these are shared across scenarios, with each scenario consisting of 100 simulation runs for each network. Each network consists of all partnerships formed over 30 years, with partnerships formed in the first two years constituting a model burn-in period (and not used in transmission simulations).

Table 1-1: Parameters that are common across all simulations

| **Parameter** | **Value(s)** | **Notes** |
| --- | --- | --- |
| **Demographic** |  | |
| Number of individuals in the model | 104 000 | Estimated number of MSM in NSW, based on the number of males in NSW in 2022 (1), with approximately 4% of males identifying as gay, bisexual, or other sexual minority orientation, and 71% of these being sexually active (2, 3). |
| **Partnership seeking behaviour** |  | |
| Seek regular partnerships only | 33% | (4, 5) |
| Seek casual partnerships only | 26% |  |
| **Number of partners in last 12 months** |  |  |
| 0 | 18.1% | (3) |
| 1 | 32.0% |  |
| 2-9 | 6.3% |  |
| 10-49 | 18.0% |  |
| 50+ | 25.6% |  |
| **Duration of a regular partnership** | 4 years on average | (6) |
| **Frequency of act** |  |  |
| Anal/oral | 4 times per week | (7) |
| Rimming | 1.5 times per week |  |
| Kissing | Assume 1.5 times the rate of oral sex. |  |
| **Risk group dependent – testing rate** |  | |
| HIV+ | 25.7% test 3+ annually, the rest test annually | (8) |
| HIV- PrEP user | 58.6% test 3+ annually, the rest test annually | (8) |
| HIV- Non-PrEP user | 7.5% test 3+ annually, 37.5% test less than once annually | (7, 8) |
| Low risk/infrequent testing | 37.5% test less than once annually | (7) |
| **Test-seeking delay upon emergence of symptom** | Mean = 2.58 days, SD = 1.49 days | (7) |

##### Sexual behaviour within a partnership

In the model, STIs can be transmitted through various types of sexual activity within partnerships. We modelled four main types of sexual activity: anal sex, oral sex, rimming, and kissing. The frequency of the different types of sexual activity considered in the model are given in Table A 2-1. Note that we define sexual activity based on the frequency of transmission between anatomical sites, which is similar but not necessarily identical to the number of acts or contacts between these sites. For example, using saliva for lubrication on a sex toy can be considered an act that leads to the transmission of STIs from the pharynx to the rectum, and is therefore interpreted as rimming in the model, even if the act of rimming never actually occurred in the modelled partnership.

Data on sexual activity is relatively scarce, necessitating broad assumptions. However, during model calibration of STI transmission parameters, variations in behaviour are likely compensated for by adjustments in transmission probabilities. For example, a decrease in transmission from oral sex could be achieved by reducing either the frequency of oral sex or the transmission probability for transmission between the penis and pharynx.

##### STI testing

In this model, individuals are tested for STIs either through participation in STI screening (asymptomatic individuals) or when actively seeking testing upon experiencing symptoms.

The model considers four STI screening risk groups: HIV-positive; HIV-negative PrEP user; HIV-negative non-PrEP user; and low-risk/infrequent testing. Individuals in each group are screened at rates as shown in . Risk groups, in turn, are determined by the number of casual partnerships an individual has in 12 months, and individuals are assigned to groups such that the STI testing behaviour for each group reflects the distribution of casual partnerships reported in Chan et al. (23). In the model, this led to 6% of the population in the HIV-positive group, 17% in the HIV-negative PrEP user group, 40% in the HIV-negative non-PrEP user group and 37% in the low-risk/infrequent testing group.

Alternatively, individuals can seek STI testing if symptoms emerge. We assume individuals will seek STI testing if they experience symptoms from any infection at any anatomical site. Consequently, even asymptomatic infections might be detected due to symptoms from other STIs and/or anatomical sites.

##### Natural history and transmission of STIs

Accurate estimates for the transmissibility and natural history parameters for STIs, including site-specific durations of infection, duration of immunity, and per-act transmission probabilities at each infection site, are often not available, or a wide range of values is reported in the literature.

In this model, most natural history parameters are determined through the model calibration process. Calibration is performed using a Nelder-Mead simplex optimisation algorithm (24) to minimize the sum of squared differences between observed STI incidences and those generated by the model in the absence of Doxy-PEP. The parameter values obtained through this calibration process are listed in Table 1-2, with further explanations for each STI provided below.

###### Gonorrhoea (Neisseria gonorrhoeae or NG)

Gonorrhoea is modelled as a localised infection specific to anatomical sites, where transmission between individuals only occurs during corresponding sexual acts when an infectious site encounters a susceptible site (e.g., an infected penis can only transmit to a susceptible rectum during anal sex). Individuals can have multiple NG infections at different anatomical sites, but an infection cannot spread between these sites without sexual contact with another partner.

NG infections are characterized by a brief exposure and immunity period. Following exposure, the affected anatomical site transitions into a non-infectious latent phase. Following a brief latency period (assumed to be 4 days), exposed sites become infectious with or without symptoms. An NG infection eventually resolves through either spontaneous clearance or treatment. After spontaneous clearance, the infected site enters a brief period of immunity (assumed to be 7 days) before becoming susceptible again. Treatment results in clearance of infection at all infected sites, making them susceptible to reinfection immediately. The course of infection is illustrated schematically in Figure S1-1.

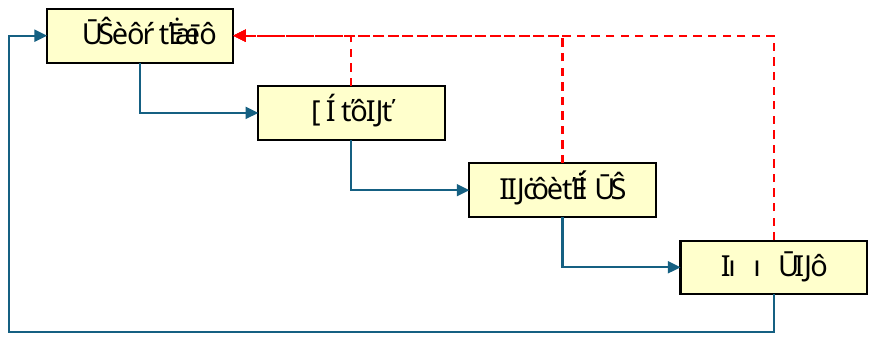

Figure S1-1: Schematic illustration of the course of infection for gonorrhoea and chlamydia in the model. The red dotted line represents the effects of treatment.

The model calibration process involves parameters that describe anatomical site-specific transmission rates, durations of infection, and the probability of symptomatic infection. Initial values (priors) for the parameters are based on values reported in published sources and previous models (22, 25, 26, 27, 28, 29, 30, 31, 32, 33). The aim of calibration is to adjust the parameters such that NG incidence from the model falls within the range of 23.9-31.5 per 100 person-years as observed in the 2023 Annual Surveillance Report.

###### Chlamydia (Chlamydia trachomatis or CT)

The natural history of chlamydia is captured in the model using the same structure as for gonorrhoea, with localised infection specific to anatomical sites and short exposure and immunity periods (see also Figure S1-1). Compared to NG infection, CT infection is thought to have a lower probability of transmission per sexual act, longer duration of infection, and a lower probability of symptomatic infection (25, 34, 35, 36). Therefore, the priors used in CT calibrations are adjusted accordingly.

Despite the more extensive published literature on the natural history of chlamydia compared to gonorrhoea, data on anatomical site-specific infections, particularly among MSM, are limited. This may be due in part to the presumed limited role of pharyngeal CT in transmission (37, 38, 39). Given that transmission of CT is likely dominated by the penile-rectal pathway, we have simplified the calibration by assuming pharyngeal CT does not contribute to transmission.

The calibration process is used to adjust the parameters such that CT incidence from the model falls within the range of 29.2-40.9 per 100 person-years as observed in the 2023 Annual Surveillance Report (13).

###### Syphilis (Treponema pallidum or TP)

Compared to gonorrhoea and chlamydia, syphilis has a more complex natural history with multiple stages and the potential to spread to various tissues throughout the body (40, 41, 42, 43). Due to this complexity and the lack of anatomical site-specific data, syphilis is modelled here as an individual-specific infection, where transmission occurs during sexual contact between individuals, rather than between specific anatomical sites.

TP infections are modelled as a multi-stage process, with different transition rates for each stage, as assumed in previous models (41, 44). Figure S1-2 is a schematic diagram that details the stages and disease progression of syphilis in this model.

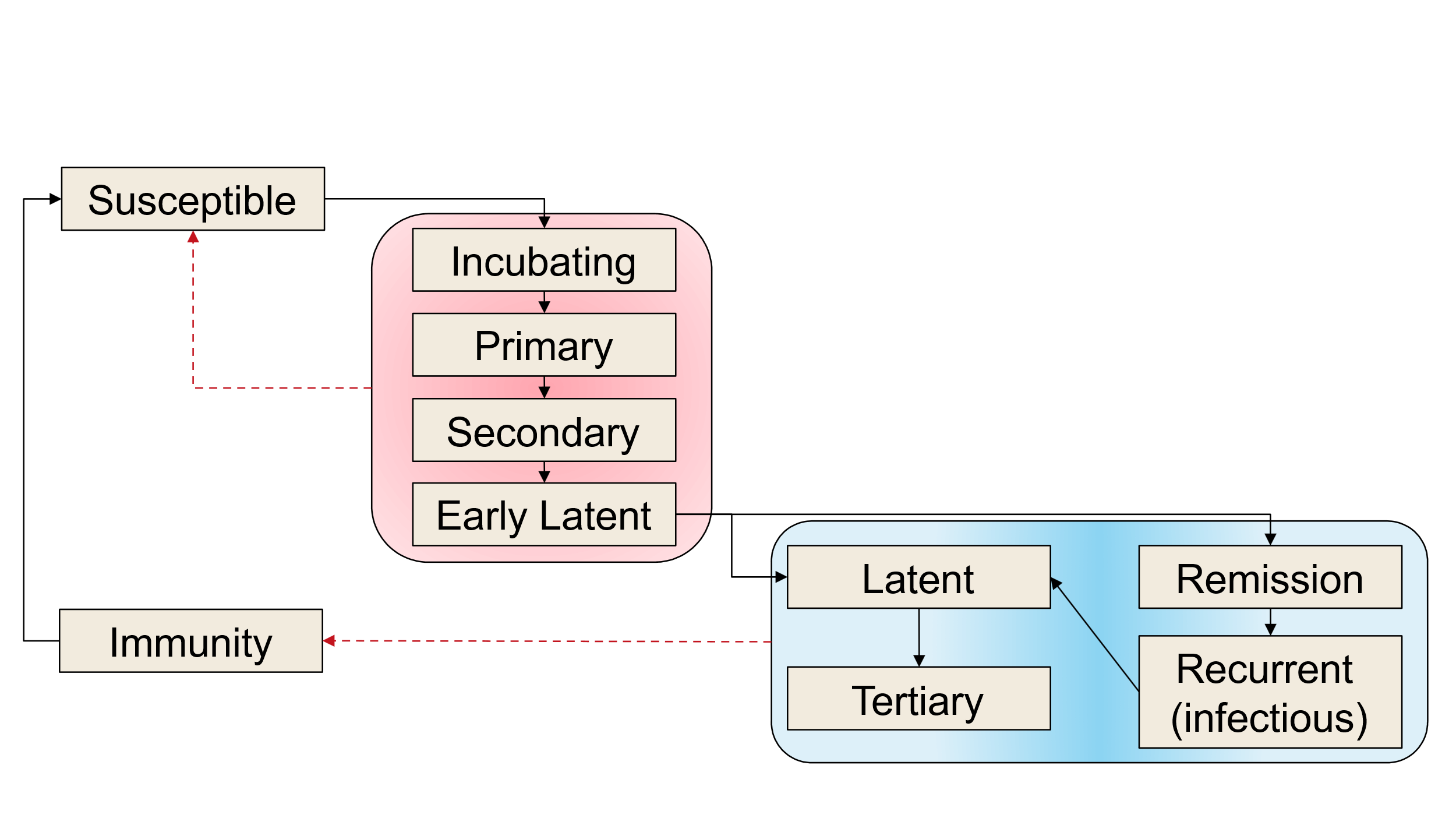

Figure S1-2: Schematic illustration of the course of syphilis infection in the model (adapted from (41), figure 1). Infected individuals are considered infectious if they are in the exposed, primary, secondary, early latent or recurrent stages of syphilis. The red dotted line represents the effects of treatment on the different stages of syphilis infection.

Due to a large number of stages and uncertainty, calibration of duration is only conducted for the primary infection stage. Durations of other infectious stages are adjusted based on the relative differences defined in previous models (41, 44). The calibration process is used to adjust the parameters such that TP incidence from the model falls within the range of 4.8-7.1 per 100 person-years as observed in the 2023 Annual Surveillance Report (13).

##### Doxy-PEP efficacy

The introduction of Doxy-PEP as an STI prevention measure is relatively recent and there is limited data on its efficacy. The preventive efficacy used in this model is based on the ANRS IPERGAY trial, which found that efficacy was around 77% for both syphilis and chlamydia (6).

The protective efficiency for gonorrhoea has been found to be lower, at around 22%, in trials (4, 5, 6). However, it is possible the efficacy for NG is even lower than that due to the high level of tetracycline resistance observed in Australian gonococcal isolates collected for antimicrobial surveillance (15).

Table 1-2: STI specific parameters (all are calibrated apart from Doxy-PEP efficacy)

|  | **TP** | **NG** | **CT** |
| --- | --- | --- | --- |
| **Transmission per act** | Incubating, primary, secondary, and recurrent stage \| 0.019; Early latent stage \| Assume to be 50% of the primary stage above (9) | Urethra to rectum \| 0.549; Rectum to urethra \| 0.339; Urethra to oropharynx \| 0.231; Oropharynx to urethra \| 0.067; Rectum to oropharynx \| 0.029; Oropharynx to rectum \| 0.013; Oropharynx to oropharynx \| 0.004 | Urethra to rectum \| 0.129; Rectum to urethra \| 0.049; Urethra to oropharynx \| 0.010; Oropharynx to urethra \| 0; Rectum to oropharynx \| 0; Oropharynx to rectum \| 0; Oropharynx to oropharynx \| 0 |
| **Duration in days if untreated** | Infectious stages (incubating, primary and secondary) \| 747-1026; Early Latent \| 594-2493; Latent \| 5475; Remission \| 180; Tertiary \| Infinity; Recurrent \| 189 | Urethra \| 61.1; Rectum \| 154.4; Oropharynx \| 108.3 | Urethra \| 385.0; Rectum \| 618.2; Oropharynx \| 289.2 |
| **Probability of infected seeking treatment (e.g., due to symptoms)** | 0.575 | Urethra \| 0.796; Rectum \| 0.443; Oropharynx \| 0 | Urethra \| 0.08; Rectum \| 0.01; Oropharynx \| 0 |
| **Doxy-PEP efficacy in preventing infection** | 77% (10-12) | 22% (10-12) | 78% (10-12) |

#### A2. Additional results

##### Infectious syphilis incidence in the absence of Doxy-PEP

Syphilis transmission in our model between 2017-2030 under the status quo scenario is shown in Figure S2-1below. Note that the model was using data exclusively from 2017 to 2021, accounting for disruptions in STI testing due to COVID-19 restrictions between early 2020 and 2021.

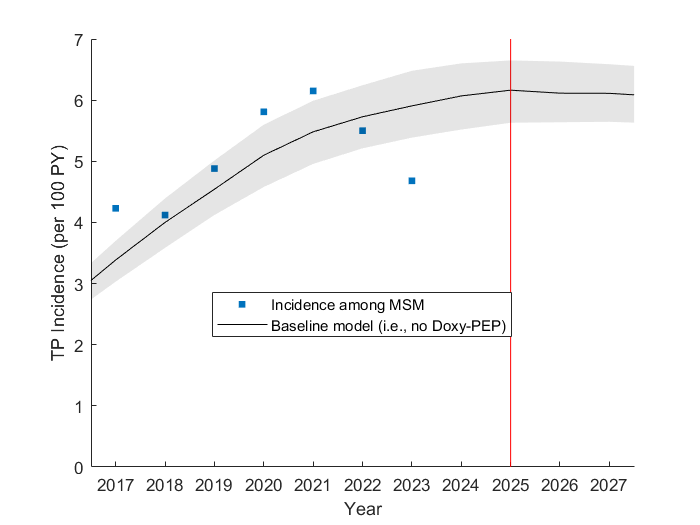

Figure S2-1: Syphilis incidence among MSM. The circle denote the adjusted syphilis incidence data from the 2024 Annual Surveillance Report, under the assumption that ~10% of MSM are HIV-positive (13). The solid line represents the median incidence from 1000 simulations of the model without Doxy-PEP, and the shaded area represents the interquartile range. The red vertical line at Year 2025 indicates the point in time when Doxy-PEP is introduced to the population for all scenarios.

##### Doxycycline consumption

Table S2-1 compares the relative reduction in syphilis incidence after five years with both the number of Doxy-PEP prescriptions issued and the doses consumed. The results suggest that approximately 36 Doxy-PEP doses per person per year are required to achieve a 50% reduction in syphilis incidence over five years. Notably, although the reductions in Scenarios 1 and 3 are similar, Scenario 3 requires fewer prescriptions per year but results in a higher proportion of Doxy-PEP doses being used during sexual contact with infectious partners. This discrepancy is driven by differences in sexual behaviour among eligible groups. For instance, individuals eligible for Doxy-PEP in Scenario 3 tend to have higher partner acquisition rates and more frequent interactions with infectious individuals, leading to more effective transmission prevention over time while requiring less Doxy-PEP.

Table S2‑1 The relationship between reduction in syphilis incidence and doxycycline consumption after 5 years of Doxy-PEP implementation (equivalent to the year 2030).

| # | Doxy-PEP eligibility | Relative reduction in syphilis incidence% (IQR%) | Doxy-PEP prescriptions per 100 person-years | Doxy-PEP doses consumed per 100 person-years, for …. | |
| --- | --- | --- | --- | --- | --- |
|  |  |  |  | **Any sexual contact** | **Sexual contact with infectious partners only** |
| 1 | Living with HIV or currently an HIV PrEP user | 53.2 (51.7 - 54.7) | 78.2 (77.1 – 79.3) | 3628.8 (3608.5-3649.1) | 449.3 (432.8-465.6) |
| 2 | Have a positive TP diagnosis | 29.7 (27.5 - 31.7) | 4.9 (4.6 – 5.3) | 1404.2 (1323.0-1483.0) | 403.0 (375.9-427.7) |
| 3 | Have a positive STI diagnosis and have had at least 1 more positive STI diagnosis in the last 12 months | 50.6 (48.1 - 53.0) | 20.8 (18.5 – 22.9) | 3600.3 (3428.8-3727.5) | 658.0 (627.9-686.5) |
| 4 | Combined effect of scenario #1, #2 and #3 | 66.7 (65.5 - 67.8) | 85.7 (84.0 – 87.5) | 5446.8 (5371.1-5510.5) | 612.3 (586.8-638.5) |

##### Effects of reduced duration, adherence, and uptake

For Scenario 3 (eligibility based on an STI diagnosis), we also consider the impact of Doxy-PEP on STI incidence when:

- Duration of use (the duration of time an individual continues to use Doxy-PEP following receipt of a prescription) is reduced from 180 days to 90 days.
- Adherence (the probability that an individual will use Doxy-PEP following a sexual encounter, as a percentage of sexual encounters), is reduced from 100% to 75%.
- Uptake Rate (the percentage of eligible individuals who commence Doxy-PEP upon STI diagnosis) is reduced from 100% to 50% or 25%.

###### STI incidence under reduced duration of use and adherence

For our baseline analysis, we assumed all individuals prescribed Doxy-PEP would use it following all sexual contacts for an average of 180 days. Table 2-1 show the findings for more realistic scenarios where these assumptions are relaxed. Here we use Scenario 3 to define eligibility for Doxy-PEP (Doxy-PEP is available to those with more than one STI diagnosis in the last 12 months) and we consider the case where the duration of use is reduced by 50% to an average of 90 days, and adherence is reduced so that Doxy-PEP is only used following 75% of sexual encounters. While reducing the duration of use (from 180 to 90 days) and adherence (from 100% to 75%) does lead to a smaller reduction in syphilis incidence (with the mean syphilis incidence decreasing from 5.9 to 2.7 per 100 PY (compared to a decrease from 5.9 to 2.3 per 100 PY for the baseline analysis), the overall impact is relatively minor.

Table S2-2: Estimated reduction in STI incidence at 5 and 10 years following the introduction of Doxy-PEP with sub-optimal use. Here, Doxy-PEP eligibility is based on STI diagnosis, with an uptake rate of 100%, and under different combinations of duration of use and adherence. The values represent the median and the interquartile range (in brackets) of the relative reduction in incidence for each STI compared to the status quo (i.e., without Doxy-PEP).

| *At 5^th^ year* | | | |
| --- | --- | --- | --- |
| Duration of Doxy-PEP use and adherence | **Syphilis %**  **(IQR%)** | **Gonorrhoea % (IQR%)** | **Chlamydia % (IQR%)** |
| Duration of use of 180 days, adherence at 100% | 50.6 (48.1 - 53.0) | 41.3 (38.3 - 45.0) | 41.3 (39.2 - 43.2) |
| Duration of use of 90 days, adherence at 100% | 41.5 (38.8 - 43.9) | 35.0 (32.0 - 38.2) | 31.5 (29.9 - 33.1) |
| Duration of use of 180 days, adherence at 75% | 43.1 (40.7 - 45.4) | 33.8 (30.8 - 37.1) | 32.3 (30.6 - 33.7) |
| Duration of use of 90 days, adherence at 75% | 34.4 (31.8 - 36.6) | 27.8 (24.6 - 31.0) | 24.2 (22.9 - 25.6) |
| *At 10^th^ year* | | | |
| Duration of Doxy-PEP use and adherence | **Syphilis %**  **(IQR%)** | **Gonorrhoea % (IQR%)** | **Chlamydia % (IQR%)** |
| Duration of use of 180 days, adherence at 100% | 57.1 (54.5 - 59.5) | 49.0 (45.5 - 53.4) | 41.7 (39.4 - 43.9) |
| Duration of use of 90 days, adherence at 100% | 47.8 (44.9 - 50.4) | 41.5 (38.2 - 45.7) | 32.2 (30.4 - 33.9) |
| Duration of use of 180 days, adherence at 75% | 50.1 (47.3 - 52.6) | 40.6 (37.2 - 44.9) | 33.3 (31.5 - 35.1) |
| Duration of use of 90 days, adherence at 75% | 40.7 (37.8 - 43.1) | 33.6 (30.4 - 37.6) | 25.2 (23.8 - 26.6) |

###### STI incidence under lower and diminishing Doxy-PEP uptake rates

The results in Table 2-2 show the impact on STI incidence if Doxy-PEP eligibility is as defined for Scenario 3 (available to those with more than one STI diagnosis in the last 12 months), but with an uptake rate (the probability of an eligible individual commencing Doxy-PEP at the time of STI diagnosis) of 100%, and reduced uptake rates of 50%, and 25%. The results suggest that even with an uptake rate as low as 25%, a 28% reduction in syphilis incidence (with the mean incidence decreasing from 5.9 to 5.0 per 100 PY) can still be achieved if Doxy-PEP is available for 5 years. Reductions of a lower magnitude are also observed for gonorrhoea and chlamydia (20% for gonorrhoea and 19% for chlamydia, respectively).

Table S2-3: Estimated reduction in STIs incidence at 5 and 10 years following the introduction of Doxy-PEP with sub-optimal uptake. Here, Doxy-PEP eligibility is based on STI diagnosis, with uptake rates of 100%, 50%, and 25%. The values represent the median and the interquartile range (in brackets) of the relative reduction in incidence for each STI compared to the status quo (i.e., without Doxy-PEP).

| *At 5^th^ year* | | | |
| --- | --- | --- | --- |
| Doxy-PEP uptake per STI diagnosis (for those who are eligible) | **Syphilis %**  **(IQR%)** | **Gonorrhoea % (IQR%)** | **Chlamydia % (IQR%)** |
| 100% | 50.6 (48.1 - 53.0) | 41.3 (38.3 - 45.0) | 41.3 (39.2 - 43.2) |
| 50% | 39.3 (36.6 - 41.9) | 30.7 (27.4 - 33.9) | 29.7 (28.0 - 31.5) |
| 25% | 27.5 (24.6 - 30.2) | 20.2 (16.9 - 23.6) | 19.4 (18.2 - 20.7) |
| *At 10^th^ year* | | | |
| Doxy-PEP uptake per STI diagnosis (for those who are eligible) | **Syphilis %**  **(IQR%)** | **Gonorrhoea % (IQR%)** | **Chlamydia% (IQR%)** |
| 100% | 57.1 (54.5 - 59.5) | 49.0 (45.5 - 53.4) | 41.7 (39.4 - 43.9) |
| 50% | 46.1 (43.1 - 48.9) | 37.0 (33.9 - 40.8) | 30.9 (28.9 - 32.8) |
| 25% | 33.5 (30.6 - 36.6) | 24.9 (21.9 - 28.4) | 20.7 (19.4 - 22.1) |

It's important to note that uptake rate in the model refers to the average probability of an eligible person accepting and commencing Doxy-PEP at the time of an STI diagnosis. This is different from an alternative implementation whereby a proportion of the population will always accept and commence Doxy-PEP and the remainer will never do so. Therefore, even if the overall rate is low, frequent testing can significantly increase an individual's probability of taking Doxy-PEP over time. For example, under a 25% uptake rate, an individual who tests annually has a greater than 75% chance of commencing Doxy-PEP at least once within 5 years.

While real-world Doxy-PEP uptake in NSW remains unknown, it is unlikely to remain constant over time. We therefore considered the possibility of uptake diminishing over time. For the results shown in Table 2-3, we assume eligibility criteria as defined for Scenario 3 with an initial uptake rate of 100%. However, the probability of uptake for each individual is reduced from 100% by 25%, 50%, or 75% each time they are offered a Doxy-PEP prescription (denoted as Doxy-PEP Rx). For example, if the reduction is 25% per Doxy-PEP prescription then the probability of using Doxy-PEP at each subsequent offer will be 1, 0.75, 0.56, 0.42, etc. The results show smaller reductions in STI incidence. In particular, to achieve a reduction of more than 40% in syphilis incidence within 5 years, more than 75% of individuals who completed their prescribed course of Doxy-PEP need to renew their prescription if they remain eligible. We note that in this case, the reduction in uptake is applied to every individual in the modelled population.

Table S2-4: Estimated reduction in STIs incidence at 5 and 10 years following the introduction of Doxy-PEP. Here, Doxy-PEP eligibility is based on STI diagnosis, with an initial uptake rate of 100% that diminishes with subsequent Doxy-PEP prescriptions (Doxy-PEP Rx.). The values represent the median and the interquartile range (in brackets) of the relative reduction in incidence for each STI compared to the status quo (i.e., without Doxy-PEP).

| *At 5^th^ year* | | | |
| --- | --- | --- | --- |
| Doxy-PEP uptake per STI diagnosis (for those who are eligible) | **Syphilis % (IQR%)** | **Gonorrhoea % (IQR%)** | **Chlamydia % (IQR%)** |
| Maintained at 100% | 50.6 (48.1 - 53.0) | 41.3 (38.3 - 45.0) | 41.3 (39.2 - 43.2) |
| Reduced by 25% per Doxy-PEP Rx. | 43.5 (41.3 - 45.7) | 33.7 (30.6 - 37.6) | 32.9 (31.6 - 33.9) |
| Reduced by 50% per Doxy-PEP Rx. | 36.0 (33.8 - 38.2) | 26.9 (23.5 - 31.2) | 25.6 (24.8 - 26.3) |
| Reduced by 75% per Doxy-PEP Rx. | 31.1 (28.7 - 33.2) | 22.4 (18.7 - 26.8) | 20.9 (20.3 - 21.6) |
| *At 10^th^ year* | | | |
| Doxy-PEP uptake per STI diagnosis (for those who are eligible) | **Syphilis % (IQR%)** | **Gonorrhoea % (IQR%)** | **Chlamydia % (IQR%)** |
| Maintained at 100% | 57.1 (54.5 - 59.5) | 49.0 (45.5 - 53.4) | 41.7 (39.4 - 43.9) |
| Reduced by 25% per Doxy-PEP Rx. | 42.9 (40.7 - 45.0) | 32.3 (28.3 - 37.5) | 26.3 (25.6 - 27.0) |
| Reduced by 50% per Doxy-PEP Rx. | 31.2 (28.8 - 33.6) | 21.7 (17.9 - 26.4) | 17.4 (17.0 - 17.9) |
| Reduced by 75% per Doxy-PEP Rx. | 25.1 (22.6 - 27.5) | 16.7 (13.4 - 20.8) | 13.3 (12.9 - 13.8) |

### References

1.	The Kirby Institute. Sexually transmissible infections are on the rise in Australia, with syphilis rates tripling over the decade. 2023 [Available from: https://www.kirby.unsw.edu.au/news/sexually-transmissible-infections-are-rise-australia-syphilis-rates-tripling-over-decade.

2.	The Kirby Institute. HIV, viral hepatitis and sexually transmissible infections in Australia: Annual surveillance report 2023. 2023 [Available from: https://www.kirby.unsw.edu.au/research/reports/asr2023.

3.	NSW Health. NSW Sexually Transmissible Infections Data Report - January to December 2023. 2024.

4.	Sokoll PR, Migliavaca CB, Doring S, Traub U, Stark K, Sardeli AV. Efficacy of postexposure prophylaxis with doxycycline (Doxy-PEP) in reducing sexually transmitted infections: a systematic review and meta-analysis. Sex Transm Infect. 2024.

5.	Luetkemeyer AF, Donnell D, Dombrowski JC, Cohen S, Grabow C, Brown CE, et al. Postexposure Doxycycline to Prevent Bacterial Sexually Transmitted Infections. N Engl J Med. 2023;388(14):1296-306.

6.	Molina JM, Charreau I, Chidiac C, Pialoux G, Cua E, Delaugerre C, et al. Post-exposure prophylaxis with doxycycline to prevent sexually transmitted infections in men who have sex with men: an open-label randomised substudy of the ANRS IPERGAY trial. Lancet Infect Dis. 2018;18(3):308-17.

7.	Cannon CA, Celum CL. Doxycycline postexposure prophylaxis for prevention of sexually transmitted infections. Topics in antiviral medicine. 2023;31(5):566-75.

8.	Cornelisse VJ, Ong JJ, Ryder N, Ooi C, Wong A, Kenchington P, et al. Interim position statement on doxycycline post-exposure prophylaxis (Doxy-PEP) for the prevention of bacterial sexually transmissible infections in Australia and Aotearoa New Zealand - the Australasian Society for HIV, Viral Hepatitis and Sexual Health Medicine (ASHM). Sex Health. 2023.

9.	New South Wales Health Centre for Population Health. NSW Sexually Transmissible Infections Strategy 2022-2026. 2022 [Available from: https://www.health.nsw.gov.au/sexualhealth/Pages/nsw-sti-strategy.aspx.

10.	Second Australian Study of Health and Relationships (ASHR2). Sex Health. 2014;11(5):381-509.

11.	GBQ+ Community Periodic Surveys. [Available from: https://www.unsw.edu.au/research/csrh/our-projects/gay-community-periodic-surveys.

12.	Traeger MW, Mayer KH, Krakower DS, Gitin S, Jenness SM, Marcus JL. Potential impact of doxycycline post-exposure prophylaxis prescribing strategies on incidence of bacterial sexually transmitted infections. Clin Infect Dis. 2023.

13.	King J, McManus H, Kwon A, Gray R, McGregor S. HIV, viral hepatitis and sexually transmissible infections in Australia: Annual surveillance report 2023. Sydney: Kirby Insitute, UNSW Sydney; 2023.

14.	Traeger MW, Guy R, Taunton C, Chow EPF, Asselin J, Carter A, et al. Syphilis testing, incidence, and reinfection among gay and bisexual men in Australia over a decade spanning HIV PrEP implementation: an analysis of surveillance data from 2012 to 2022. The Lancet Regional Health – Western Pacific. 2024;51.

15.	Cornelisse VJ, Riley B, Medland NA. Australian consensus statement on doxycycline post-exposure prophylaxis (doxy-PEP) for the prevention of syphilis, chlamydia and gonorrhoea among gay, bisexual and other men who have sex with men. Med J Aust. 2024;220(7):381-6.

16.	Australian Bureau of Statistics. National, state and territory population Canberra: ABS; 2023 [Available from: https://www.abs.gov.au/statistics/people/population/national-state-and-territory-population/jun-2023.

17.	Wilson T, Temple J, Lyons A, Shalley F. What is the size of Australia's sexual minority population? BMC Res Notes. 2020;13(1):535.

18.	Grulich AE, de Visser RO, Badcock PB, Smith AMA, Heywood W, Richters J, et al. Homosexual experience and recent homosexual encounters: the Second Australian Study of Health and Relationships. Sexual health. 2014;11(5):439-50.

19.	Lee E, Mao L, Broady T, Bavinton B, McKenzie T, Batrouney C, et al. Gay Community Periodic Survey: Melbourne 2018. Sydney: Centre for Social Research in Health, UNSW Sydney; 2018.

20.	Broady T, Chan C, Bavinton B, Mao L, Molyneux A, Delhomme F, et al. Gay Community Periodic Survey: Sydney 2021. Sydney: Centre for Social Research in Health, UNSW Sydney; 2021.

21.	Prestage GP, Hudson J, Bradley J, Down I, Sutherland J, Corrigan N, et al. TOMS - Three or More Study. Sydney: National Centre in HIV Epidemiology and Clinical Research, University of New South Wales; 2008.

22.	Duan Q, Carmody C, Donovan B, Guy RJ, Hui BB, Kaldor JM, et al. Modelling response strategies for controlling gonorrhoea outbreaks in men who have sex with men in Australia. PLoS Comput Biol. 2021;17(11):e1009385.

23.	Chan C, Holt M, Broady TR, Traeger MW, Mao L, Grulich AE, et al. Trends in Testing and Self-Reported Diagnoses of Sexually Transmitted Infections in Gay and Bisexual Men in Australia, 2017 to 2021: Analysis of National Behavioral Surveillance Surveys. Sex Transm Dis. 2023;50(12):789-95.

24.	Lagarias JC, Reeds JA, Wright MH, Wright PE. Convergence properties of the Nelder-Mead simplex method in low dimensions. Siam J Optimiz. 1998;9(1):112-47.

25.	Johnson LF, Alkema L, Dorrington RE. A Bayesian approach to uncertainty analysis of sexually transmitted infection models. Sexually Transmitted Infections. 2010;86:169-74.

26.	Garnett GP, Mertz KJ, Finelli L, Levine WC, St Louis ME. The transmission dynamics of gonorrhoea: modelling the reported behaviour of infected patients from Newark, New Jersey. Philosophical Transactions: Biological Sciences. 1999;354(1384):787-97.

27.	Hui BB, Padeniya TN, Rebuli N, Gray RT, Wood JG, Donovan B, et al. A Gonococcal Vaccine Has the Potential to Rapidly Reduce the Incidence of Neisseria gonorrhoeae Infection Among Urban Men Who Have Sex With Men. J Infect Dis. 2022;225(6):983-93.

28.	Barbee LA, Soge OO, Khosropour CM, Haglund M, Yeung W, Hughes J, et al. The Duration of Pharyngeal Gonorrhea: A Natural History Study. Clin Infect Dis. 2021.

29.	Unemo M, Seifert HS, Hook EW, 3rd, Hawkes S, Ndowa F, Dillon JR. Gonorrhoea. Nat Rev Dis Primers. 2019;5(1):79.

30.	Bissessor M, Tabrizi SN, Fairley CK, Danielewski J, Whitton B, Bird S, et al. Differing Neisseria gonorrhoeae bacterial loads in the pharynx and rectum in men who have sex with men: implications for gonococcal detection, transmission, and control. J Clin Microbiol. 2011;49(12):4304-6.

31.	Brunham RC, Garnett GP, Swinton J, Anderson RM. Gonococcal infection and human fertility in sub-Saharan Africa. Proceedings of the Royal Society London B: Biological Sciences. 1991;246(1316):173-7.

32.	Fairley CK, Chen MY, Bradshaw CS, Tabrizi SN. Is it time to move to nucleic acid amplification tests screening for pharyngeal and rectal gonorrhoea in men who have sex with men to improve gonorrhoea control? Sexual health. 2011;8(1):9-11.

33.	Ong JJ, Fethers K, Howden BP, Fairley CK, Chow EPF, Williamson DA, et al. Asymptomatic and symptomatic urethral gonorrhoea in men who have sex with men attending a sexual health service. Clin Microbiol Infect. 2017;23(8):555-9.

34.	Rönn MM, Wolf EE, Chesson H, Menzies NA, Galer K, Gorwitz R, et al. The Use of Mathematical Models of Chlamydia Transmission to Address Public Health Policy Questions: A Systematic Review. Sexually Transmitted Diseases. 2017;44(5):278-83.

35.	Barbee LA, Khosropour CM, Dombrowski JC, Manhart LE, Golden MR. An estimate of the proportion of symptomatic gonococcal, chlamydial and non-gonococcal non-chlamydial urethritis attributable to oral sex among men who have sex with men: a case-control study. Sex Transm Infect. 2015.

36.	Althaus CL, Heijne JC, Low N. Towards more robust estimates of the transmissibility of Chlamydia trachomatis. Sex Transm Dis. 2012;39(5):402-4.

37.	Evers YJ, Dukers-Muijrers N, van Liere G, van Bergen J, Kuizenga-Wessel S, Hoebe C. Pharyngeal Chlamydia trachomatis in Men Who Have Sex With Men (MSM) in The Netherlands: A Large Retrospective Cohort Study. Clin Infect Dis. 2022;74(8):1480-4.

38.	Phillips TR, Fairley CK, Maddaford K, Danielewski J, Hocking JS, Lee D, et al. Bacterial Load of Chlamydia trachomatis in the Posterior Oropharynx, Tonsillar Fossae, and Saliva among Men Who Have Sex with Men with Untreated Oropharyngeal Chlamydia. J Clin Microbiol. 2019;58(1).

39.	Chow EP, Fairley CK. The role of saliva in gonorrhoea and chlamydia transmission to extragenital sites among men who have sex with men: new insights into transmission. J Int AIDS Soc. 2019;22 Suppl 6(Suppl Suppl 6):e25354.

40.	Lafond RE, Lukehart SA. Biological basis for syphilis. Clin Microbiol Rev. 2006;19(1):29-49.

41.	National Centre in HIV Epidemiology and Clinical Research. Phase A of the National Gay Men’s Syphilis Action Plan: Modelling evidence and research on acceptability of interventions for controlling syphilis in Australia. Sydney: National Centre in HIV Epidemiology and Clinical Research; 2009.

42.	Garnett GP, Aral SO, Hoyle DV, Cates W, Jr., Anderson RM. The natural history of syphilis. Implications for the transmission dynamics and control of infection. Sex Transm Dis. 1997;24(4):185-200.

43.	Gurney Clark E, Danbolt N. The Oslo Study of the Natural Course of Untreated Syphilis: An Epidemiologic Investigation Based on a Re-study of the Boeck-Bruusgaard Material. Medical Clinics of North America. 1964;48(3):613-23.

44.	Hui BB, Ward JS, Guy R, Law MG, Gray RT, Regan DG. Impact of Testing Strategies to Combat a Major Syphilis Outbreak Among Australian Aboriginal and Torres Strait Islander Peoples: A Mathematical Modeling Study. Open Forum Infect Dis. 2022;9(5):ofac119.

1.	Australian Bureau of Statistics. National, state and territory population Canberra: ABS; 2023 [Available from: <https://www.abs.gov.au/statistics/people/population/national-state-and-territory-population/jun-2023>.

2.	Wilson T, Temple J, Lyons A, Shalley F. What is the size of Australia's sexual minority population? BMC Res Notes. 2020;13(1):535.

3.	Grulich AE, de Visser RO, Badcock PB, Smith AMA, Heywood W, Richters J, et al. Homosexual experience and recent homosexual encounters: the Second Australian Study of Health and Relationships. Sexual health. 2014;11(5):439-50.

4.	Lee E, Mao L, Broady T, Bavinton B, McKenzie T, Batrouney C, et al. Gay Community Periodic Survey: Melbourne 2018. Sydney: Centre for Social Research in Health, UNSW Sydney; 2018.

5.	Broady T, Chan C, Bavinton B, Mao L, Molyneux A, Delhomme F, et al. Gay Community Periodic Survey: Sydney 2021. Sydney: Centre for Social Research in Health, UNSW Sydney; 2021.

6.	Prestage GP, Hudson J, Bradley J, Down I, Sutherland J, Corrigan N, et al. TOMS - Three or More Study. Sydney: National Centre in HIV Epidemiology and Clinical Research, University of New South Wales; 2008.

7.	Duan Q, Carmody C, Donovan B, Guy RJ, Hui BB, Kaldor JM, et al. Modelling response strategies for controlling gonorrhoea outbreaks in men who have sex with men in Australia. PLoS Comput Biol. 2021;17(11):e1009385.

8.	Chan C, Holt M, Broady TR, Traeger MW, Mao L, Grulich AE, et al. Trends in Testing and Self-Reported Diagnoses of Sexually Transmitted Infections in Gay and Bisexual Men in Australia, 2017 to 2021: Analysis of National Behavioral Surveillance Surveys. Sex Transm Dis. 2023;50(12):789-95.

9.	National Centre in HIV Epidemiology and Clinical Research. Phase A of the National Gay Men’s Syphilis Action Plan: Modelling evidence and research on acceptability of interventions for controlling syphilis in Australia. Sydney: National Centre in HIV Epidemiology and Clinical Research; 2009.

10.	Sokoll PR, Migliavaca CB, Doring S, Traub U, Stark K, Sardeli AV. Efficacy of postexposure prophylaxis with doxycycline (Doxy-PEP) in reducing sexually transmitted infections: a systematic review and meta-analysis. Sex Transm Infect. 2024.

11.	Luetkemeyer AF, Donnell D, Dombrowski JC, Cohen S, Grabow C, Brown CE, et al. Postexposure Doxycycline to Prevent Bacterial Sexually Transmitted Infections. N Engl J Med. 2023;388(14):1296-306.

12.	Molina JM, Charreau I, Chidiac C, Pialoux G, Cua E, Delaugerre C, et al. Post-exposure prophylaxis with doxycycline to prevent sexually transmitted infections in men who have sex with men: an open-label randomised substudy of the ANRS IPERGAY trial. Lancet Infect Dis. 2018;18(3):308-17.

13.	King J, Kwon JA, McManus H, Gray R, McGregor S. HIV, viral hepatitis and sexually transmissible infections in Australia: Annual surveillance report 2024. Sydney: Kirby Insitute, UNSW Sydney; 2024.
